# Automated Intracranial Thrombus Segmentation from CT Images of Patients with Acute Ischemic Stroke: A Dual-Channel nnU-Net Approach with Uncertainty Quantification

**DOI:** 10.64898/2026.01.23.26344730

**Authors:** Tatsat R. Patel, Vinay Jaikumar, Naoki Kaneko, Amy Letavay, Maxim Mokin, Robert W. Raegenhardt, Elad I. Levy, Adnan H. Siddiqui, Vincent M. Tutino

## Abstract

**Background:** Automated thrombus segmentation on CT imaging could enable routine extraction of clot volume and other biomarkers in large vessel occlusion (LVO) stroke, but current deep learning models provide deterministic masks without indicating when their output is unreliable. We developed and evaluated an uncertainty-aware segmentation framework that couples nnU-Net with Bayesian-style uncertainty estimation to support quality-controlled use of automated clot measurements.

**Methods:** In this single-center retrospective study, we included MT-treated AIS patients with baseline NCCT and CTA. NCCT was rigidly registered to CTA, and an atlas-based pipeline cropped images to a supratentorial intracranial arterial ROI. Clots were manually segmented on co-registered NCCT/CTA. A 3D nnU-Net with two-channel input (CTA+NCCT) was trained with a cyclical learning-rate schedule; ensembles of checkpoints were used to approximate the predictive posterior and compute voxel-wise entropy. Case-level clot uncertainty (U_clot) was defined as mean entropy within the predicted clot. We assessed segmentation metrics, volumetric agreement, the relationship between U_clot and Dice, and the performance of U_clot for triaging poor segmentations (Dice <0.60).

**Results:** In the test cohort (n=80), mean Dice was 0.64±0.24 and volumetric ICC 0.83, with strong correlation between predicted and ground-truth volumes (ρ=0.77, R²=0.69). Performance was higher for hyperdense vs non-hyperdense clots and for medium/large vs small clots. U_clot was strongly inversely associated with Dice (ρ=−0.70 overall) and remained informative within all phenotype subgroups. As a univariate predictor of poor segmentation, U_clot achieved an AUC of 0.89; the optimal threshold (0.323) yielded 90% sensitivity and 96% negative predictive value, allowing 60% of cases to be accepted automatically while improving volume-category agreement from 66% (κ=0.49) to 83% (κ=0.69).

**Conclusions:** Uncertainty-aware nnU-Net segmentation provides human-level thrombus delineation while supplying a robust, interpretable case-level confidence score. Using U_clot to triage segmentations can substantially enhance the reliability of clot volume categorization, offering a practical pathway toward safe deployment of automated clot analysis in stroke care and research.

## Introduction

Acute ischemic stroke (AIS) due to large vessel occlusion (LVO) is a major cause of death and disability worldwide.^1^ Endovascular mechanical thrombectomy (MT) and intravenous tissue plasminogen activator (IV-tPA) are now standard-of-care reperfusion strategies for appropriately selected patients with LVO, with randomized trials establishing the benefit of MT over medical therapy alone in anterior circulation LVO.^2–4^ However, individual patients vary widely in their likelihood of recanalization and functional recovery with IVT, MT, or combined approaches.^4^ Thrombus location, morphology (length and volume) and density (on CT imaging, i.e. Hyper-dense artery sign, perviousness), have been repeatedly associated with recanalization after both IV alteplase and MT, as well as with functional outcome.^5–9^ More recently, clot radiomics has been shown to predict IVT success and first-pass recanalization with aspiration MT, underscoring the potential of imaging-based clot phenotyping to support treatment selection.^10–15^

Extracting these predictive features crucially depends on our ability to accurately segment the clot tissue in the medical imaging.^16–19^ Currently, thrombi on CT imaging (CT angiography [CTA] and non-contrast CT [NCCT]) are often segmented manually.^20^ This process is labor-intensive and difficult to perform quickly and reliably. Moreover, it is highly observer dependent, with only a 50–60% volumetric overlap reported between rater annotations, reflecting the small size and low contrast of thrombi on CT and highlighting the need for more standardized, automated approaches.^16–18^ Deep learning offers a robust solution for rapid and consistent clot segmentation. So far there have only been 2 feasibility studies demonstrating deep-learning clot segmentation methods.^16,17^ While limited, they reported dice coefficients (overlap between human segmentation and AI prediction) of 62% and 71% respectively in their corresponding testing cohorts.^16,17^ These findings show that deep learning can reach or even exceed human-level agreement with manual clot masks. Yet work remains to be done, as they also demonstrated that even the best models still disagree with expert segmentations in roughly 30–35% of voxels.

These previous methods only provide a single, deterministic mask for clot segmentation, with no accompanying estimate of segmentation quality. Indeed, a lack of uncertainty awareness is problematic in a setting where both data and labels are noisy.^21^ Thrombi are small structures relative to the brain, are obscured by noise and artifacts, and may not be consistently visible on NCCT or CTA depending on composition and collateral flow. Moreover, manual segmentations are themselves imperfect and inconsistent. A deterministic network trained on such data can achieve a respectable average Dice, yet still fail silently on individual cases, precisely the cases where clinicians most need to know that the output is unreliable. Recent work in other domains has therefore begun to pair semantic segmentation AI methods (like nnUNet) with explicit uncertainty quantification, for example by using cyclical learning-rate schedules and ensembles of checkpoints at different local minima to generate voxel-wise entropy maps and image-level uncertainty scores.^22,23^ To date, however, uncertainty-aware segmentation has not been applied to intracranial thrombus segmentation.

In this study, we develop an automated, uncertainty-aware framework for thrombus segmentation on CT imaging in patients with LVO who underwent MT. Building on nnU-Net and checkpoint-ensemble–based Bayesian approximation,^22^ we generate both a clot mask and an associated voxel-wise uncertainty map for each patient and summarize the latter into a case-level clot uncertainty score (U_clot). We then: (1) quantify the sensitivity of the segmentation model to clot density and size, (2) determine how uncertainty relates to standard segmentation quality metrics and propagates into clinically relevant downstream tasks such as clot volume categorization; and (3) explore a triage strategy in which high-certainty cases are processed fully automatically, while high-uncertainty cases are flagged for expert review and correction.

## Methods

### Study Participants

We conducted a retrospective, single-center study of patients with acute ischemic stroke due to large vessel occlusion (LVO) both in the anterior and posterior circulation or selected medium vessel occlusion (M2–M3) who underwent endovascular mechanical thrombectomy (MT) at our institution. All included patients had baseline non-contrast CT (NCCT) and CT angiography (CTA) acquired before MT procedures were performed. MT was performed in all cases at the discretion of the treating neurointerventionalist, with some patients also receiving intravenous thrombolysis (IVT) per standard clinical criteria. For inclusion, images were required to have thin-slice reconstructions (typically 0.4–0.6 mm; a small minority had 1.0-mm slices) and sufficient quality for analysis. We excluded cases with severe motion, beam-hardening, or other artifacts that precluded reliable clot visualization, as well as cases in which rigid registration between NCCT and CTA failed. For this retrospective analysis, the requirement for individual informed consent was waived. The study was approved by the institutional review board at the University at Buffalo (study 00002092).

### Imaging and Ground-Truth Generation

#### CT acquisition and preprocessing

Baseline NCCT and CTA were acquired as part of routine clinical stroke imaging. Most examinations were performed on a 320-detector-row scanner (Aquilion ONE, Canon Medical Systems), with a small minority obtained on comparable multidetector CT systems from other vendors. For NCCT, typical acquisition parameters were 135 kVp and 370–600 mA; for CTA, 120 kVp and 150–205 mA with a bolus of non-ionic iodinated contrast agent (350 mg I/mL) administered at 5 mL/s.^24,25^ Thin-slice reconstructions were used for all analyses. NCCT volumes had a median voxel size of 0.5 × 0.439 × 0.439 mm³, and CTA volumes were reconstructed with approximately isotropic 0.5-mm voxels.

All image preparation and manual annotations were performed in 3D Slicer (version 5.0.3).^26^ For each patient, the NCCT was rigidly co-registered and resampled onto the CTA using the BRAINSFit module, with a 7-parameter transform (rigid + global scale), Mattes mutual information as the similarity metric, and low-percentage stochastic sampling.^27^ This ensured voxel-wise alignment of the hyperdense artery sign (if visible) on NCCT with the intraluminal filling defect on CTA, and provided a common space for downstream region-of-interest (ROI) extraction and segmentation.

#### Atlas-based vascular ROI definition and cropping

Because thrombi occupy only a small fraction of the cranial volume and are confined to the proximal intracranial arterial tree, we restricted all subsequent processing to a standardized vascular ROI. We began by defining a supratentorial vascular box in a publicly available head atlas covering the major intracranial arteries of the circle of Willis and proximal branches.^28^ The ROI was manually placed to encompass the intracranial internal carotid arteries from the skull base through the supraclinoid segments, the M1–M3 segments of the middle cerebral arteries, the A1–A2 segments of the anterior cerebral arteries, the basilar artery, and at least the P1–P2 segments of the posterior cerebral arteries, with a small margin to account for anatomic variability and registration errors.

To transfer this atlas-space ROI to each patient, we used a two-stage registration strategy implemented in SimpleITK and Elastix.^29,30^ First, the NCCT was cropped to the supratentorial half to reduce non-informative inferior anatomy and improve registration robustness. An initial Euler 3D transform was estimated using CenteredTransformInitializer to roughly align the atlas to the patient NCCT, and this transform was used to resample both the atlas image and the atlas-space ROI mask into NCCT space. Second, we refined the alignment with an affine registration using Elastix (default affine parameter map with increased iterations and sampling density), again using the NCCT as the fixed image. The resulting composite transform was applied to the atlas ROI mask, which was then thresholded, morphologically closed, and reduced to its largest connected component to obtain a patient-specific intracranial vascular mask.

Finally, we computed the tight bounding box of this refined mask and used it to crop the NCCT, CTA, and (when available) clot label maps to a common patient-specific ROI. This pipeline ensured that all downstream segmentation and model training focused on the main anterior and posterior circulation arteries with a small safety margin, reducing class imbalance, memory requirements, and the risk of spurious predictions in regions far from the intracranial vasculature.

#### Manual clot segmentation

Within the cropped NCCT–CTA ROIs, ground-truth clot segmentations were generated manually by an experienced rater. Segmentation was performed primarily on axial CTA images, with sagittal and coronal reconstructions used to refine the contours. The proximal and distal limits of the thrombus were defined by the intraluminal filling defect on CTA, while the hyperdense artery sign (if present) on co-registered NCCT was used as an additional guide to identify clot boundaries in ambiguous regions. This approach mirrors our previously published clot segmentation and radiomics workflow, in which clots were segmented on co-registered NCCT and CTA and validated in multiple downstream analyses.^12,18^

To place our annotations in the context of known inter-rater variability for clot segmentation, we refer to two prior studies from our group. In an inter-user radiomics sensitivity analysis of 17 MT-treated patients, three observers independently segmented clots on NCCT and CTA; pairwise volumetric agreement yielded Dice similarity coefficients of approximately 0.56–0.60 with average surface distances of ∼1.3–1.7 voxels.^18^ More recently, in a paired CT radiomics–transcriptomics analysis of stroke thrombi from our laboratory, an inter-rater assessment of clot annotations again demonstrated high agreement, further supporting the robustness of our in-house clot segmentation workflow.^31^ Together with the published inter-user analysis above, these data indicate that the manual masks used as ground truth in the present study are representative of expert consensus and suitable for training and evaluating automated clot segmentation models.

### Deep-Learning Approach

#### Network architecture and input representation

We used the self-configuring nnU-Net framework as our base segmentation model.^23^ For the present task, nnU-Net was configured in its 3D full-resolution setting, with all hyperparameters (network depth, feature map widths, patch size, and batch size) automatically derived from the dataset “fingerprint” according to the original implementation.

Within each patient-specific vascular ROI, we constructed a two-channel input volume by stacking the cropped CTA and co-registered NCCT (ROI-CTA and ROI-NCCT) along the channel dimension. The corresponding cropped manual clot mask served as the binary ground-truth label (clot vs background). Thus, the model learned to exploit both intraluminal contrast information from CTA and density information from NCCT to delineate thrombi. All images were preprocessed using the standard nnU-Net pipeline, including intensity normalization and resampling to a task-specific target spacing.^23^ The network was trained with nnU-Net’s default combination of soft Dice and cross-entropy loss, optimized with stochastic gradient descent with momentum.

#### Cyclical learning-rate schedule and checkpoint ensemble

To obtain Bayesian-style uncertainty estimates without modifying the network architecture, we adopted the trajectory-based posterior sampling strategy of Zhao et al.^22^ and combined it with a cyclical learning-rate (LR) schedule with warm restarts, inspired by Rel-UNet and SGDR.^32^ The key idea is that, once the learning rate has decayed and the training has stabilized, successive SGD updates explore a local neighborhood of a solution in weight space;^33^ saving these checkpoints yields approximate samples from the weight posterior, which can be used to build an ensemble and quantify uncertainty.

We parameterized the learning-rate schedule as follows: the network was trained for a total of 1200 epochs, divided into 3 cycles of 400 epochs (𝑇_𝑐_) each. At the start of each cycle, the learning rate was reset to a peak value of 0.01 (𝜂_𝑚𝑎𝑥_), then decayed toward a minimum learning rate of 0.0001 (𝜂_𝑚𝑖𝑛_). Within each cycle, the learning rate was held at this minimum (“flat” phase) for approximately 20% (i.e.,𝛼 = 0.20) of the epochs, during which model weights explored a local neighborhood around a solution and checkpoints were saved. At the end of training, we retained the 10 best-performing checkpoints on the validation set for use in the ensemble-based uncertainty estimation.

During the decay phase, i.e. the first (1 − 𝛼)𝑇_𝑐_ epochs, the learning rate was scheduled using the following equation,

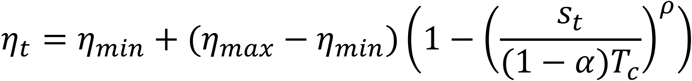

Where 𝛼 is the decay exponent as in Zhao et al., 𝑠_𝑡_ is the in-cycle epoch index, and 𝜂_𝑡_ is the in-cycle indexed learning-rate value.^22^ Following the decay phase, the flat phase (last 𝛼𝑇_𝑐_ epochs) follows a simple constant learning rate of 𝜂_𝑚𝑖𝑛_. At the start of each cycle, the LR was restarted to 𝜂_𝑚𝑎𝑥_, nudging the weights out of the previous basin and encouraging exploration of multiple local minima in weight space, similar in spirit to SGDR and multi-modal sampling in Zhao et al.^22^

#### Predictive posterior and voxel-wise entropy

Given a test input volume 𝑥 (two-channel CTA+NCCT ROI), each checkpoint 𝑤^(𝑚)^ produces a voxel-wise softmax output 𝑝(𝑦 | 𝑥, 𝑤^(𝑚)^), where 𝑦 ∈ {0,1} denotes background versus clot. The Bayesian predictive distribution is approximated by averaging over the ensemble:

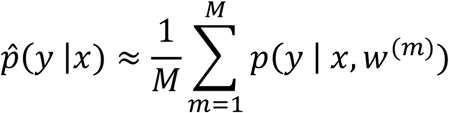

With 𝑀 = 10 in our implementation. Over each voxel, the ensemble-averaged clot probability is then defined as 𝑝̂_𝑣_ = 𝑝̂(𝑦 |𝑥, 𝑤^(𝑚)^)_𝑣_.^22^ Shannon entropy can be computed from this binary predictive distribution,

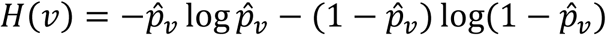

This yields a voxel-wise uncertainty map i.e. 𝐻(𝑣), which is typically highest along and around the clot boundary where the ensemble members disagree most strongly, as observed in prior work on uncertainty-aware nnU-Net segmentation.^22,32^ This map 𝐻(𝑣) is also used to visualize and qualitatively analyze areas of the clot in individual cases where the model is uncertain in its prediction.

#### Case-level clot uncertainty score

To summarize uncertainty at the case level, we define a clot-specific uncertainty index 𝑈_𝑐𝑙𝑜𝑡_ by averaging the normalized entropy over the predicted clot region. Let Ω_𝑐𝑙𝑜𝑡_ = {𝑣 |𝑀̂_𝑣_ = 1} be the set of voxels in the predicted clot. Then,

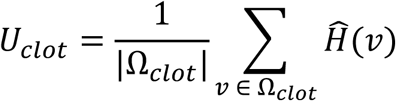

Thus, 𝑈_𝑐𝑙𝑜𝑡_ ∈ [0,1] measures the mean predictive uncertainty inside the AI-predicted clot mask. Low values indicate that all checkpoints strongly agree on both the presence and extent of the clot (high-confidence segmentation), whereas higher values indicate substantial disagreement among posterior samples, typically reflecting ambiguous clot boundaries, low contrast, or label noise.

### Evaluation Metrics and Statistical Analysis

#### Ensemble selection and final segmentation rule

For all analyses, voxel-wise mean probability, entropy, mutual information, and the U_clot score were computed from an ensemble of checkpoints obtained with the cyclical learning-rate schedule described above. Within each learning-rate cycle, we saved ten checkpoints during the flat (minimum learning-rate) phase; to balance computational cost and diversity, we retained four per cycle (the 1st, 4th, 7th, and 10th) for inference, yielding a 60-model ensemble across the three cycles per each of the 5-fold training.

For each voxel, the ensemble-mean clot probability and entropy were computed as previously described. Mutual information was defined in the standard BALD formulation as the difference between the entropy of the ensemble-mean predictive distribution and the mean entropy of the individual models, thereby isolating epistemic uncertainty.^34^ Voxels were finally classified as clot or background using a joint probability–uncertainty rule based on the ensemble-mean probability and mutual information, with empirically chosen thresholds on the validation set (e.g., retaining voxels with sufficiently high mean clot probability and sufficiently low mutual information); this composite rule was kept fixed for all test-set analyses.

#### Segmentation performance metrics

Segmentation performance was evaluated at the case level by comparing predicted clot masks with the corresponding manual ground-truth masks. We computed: Dice similarity coefficient, Precision (positive predictive value), Recall (sensitivity), Intersection-over-union (IoU), Surface Dice at 1 mm using a symmetric surface-distance tolerance of 1 mm, and, Volumetric similarity, defined as

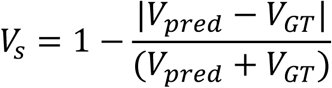

where 𝑉_𝑝𝑟𝑒𝑑_ and 𝑉_𝐺𝑇_ are predicted and ground-truth volumes respectively.

These metrics were reported for the full test cohort and separately within predefined clot subgroups. Hyperdense versus non-hyperdense thrombi were defined based on the median HU within the ground-truth clot mask on NCCT, with hyperdense clots having median HU > 51 HU.^35,36^ Clot size was categorized using the ground-truth clot volume into three groups: small (0 < V < 50 mm³), medium (50 ≤ V < 150 mm³), and large (V ≥ 150 mm³).^37,38^

#### Volumetric agreement and correlation analyses

Agreement between predicted and ground-truth clot volumes was evaluated using several complementary approaches. We generated Bland–Altman plots of 𝑉_𝑝𝑟𝑒𝑑_ − 𝑉_𝐺𝑇_ versus the mean (𝑉_𝑝𝑟𝑒𝑑_ + 𝑉_𝐺𝑇_)/2 to visualize bias and limits of agreement, and quantified association and calibration between 𝑉_𝑝𝑟𝑒𝑑_ and 𝑉_𝐺𝑇_ using correlation and simple linear regression. Volumetric agreement was further summarized with intraclass correlation coefficients (two-way mixed-effects, absolute-agreement model). All analyses were performed for the overall cohort and stratified by hyperdensity status and clot size category.

#### Uncertainty analyses

Uncertainty was assessed at both voxel and case level. For each case, we summarized the voxel-wise normalized entropy within the predicted clot region to obtain the scalar U_clot score (mean normalized entropy inside the predicted mask). We then examined the Relationship between segmentation quality and uncertainty. Dice, surface Dice at 1 mm, volumetric similarity, and recall were analyzed as functions of U_clot using correlation analyses. We performed these analyses both on the full cohort and within subgroups (hyperdense vs non-hyperdense; small, medium, large clots) to assess whether the uncertainty–performance relationship was consistent across clot phenotypes. We also tested the groupwise differences in uncertainty by comparing U_clot between hyperdense and non-hyperdense clots and across size categories. Analogous tests were performed for segmentation metrics themselves (e.g., Dice, surface Dice, recall) to evaluate whether uncertainty paralleled performance differences between groups.

#### Uncertainty-based triage analysis

To evaluate U_clot as a triage metric, we dichotomized cases into “well segmented” and “poorly segmented” using a Dice threshold of 0.60 (Dice ≥ 0.60 vs < 0.60). Treating this binary label as the reference, we used receiver operating characteristic (ROC) analysis with U_clot as a continuous predictor to assess discrimination and selected an optimal U_clot threshold using Youden’s index.

At this threshold, we summarized performance using confusion matrices, sensitivity, specificity, predictive values, accuracy, and the proportion of cases that would be automatically accepted (low U_clot) versus flagged for expert review (high U_clot). We also performed a sensitivity analysis by varying the U_clot threshold and plotting (i) the fraction of cases that would be automatically accepted and (ii) the mean Dice (and surface Dice) among accepted cases to illustrate the trade-off between automation and segmentation quality. Further details on the statistical framework for ROC analysis and decision-curve analysis are provided in the General statistical considerations section.

#### Effect of triage on categorical clot size agreement

As a downstream use case, we assessed how uncertainty-based triage affected agreement in clot size categorization, which is clinically relevant for treatment selection. For each case, ground-truth and predicted volumes were independently discretized into the three size categories (small/medium/large) defined above. We then constructed 3×3 confusion matrices comparing predicted vs ground-truth categories and computed overall classification accuracy and Cohen’s κ as measures of categorical agreement.

These analyses were first performed on the entire test cohort and then repeated after excluding high-uncertainty cases based on the optimal U_clot threshold from the ROC analysis, to quantify how uncertainty-based triage influenced categorical agreement. The statistical interpretation of accuracy and κ follows the criteria outlined in the General statistical considerations section.

#### General statistical considerations

All tests were two-sided with a significance level of α = 0.05. Continuous variables are reported as mean ± standard deviation, and categorical variables as counts and percentages. Normality was assessed using Shapiro–Wilk test, and this guided the choice of Student’s t test or one-way ANOVA versus Mann–Whitney U or Kruskal–Wallis tests for group comparisons. Associations between continuous measures (e.g., predicted vs ground-truth volume, U_clot vs Dice) were quantified using Pearson or Spearman correlation and simple linear regression (slope, intercept, R²) for calibration. Agreement in continuous measures was evaluated with Bland–Altman analysis and intraclass correlation coefficients (two-way mixed-effects, absolute-agreement model),^39^ interpreted using conventional thresholds (poor <0.50, moderate 0.50–0.75, good 0.75–0.90, excellent >0.90).^40^ Categorical agreement for volume-based size classes was summarized with accuracy and Cohen’s κ. For uncertainty-based triage, receiver operating characteristic (ROC) curves were constructed using U_clot as a continuous predictor of “well segmented” (Dice ≥ 0.60) versus “poorly segmented” cases, with area under the curve (AUC) and an optimal cut-off chosen by Youden’s index. At this cut-off, we reported confusion matrices, sensitivity, specificity, predictive values, and accuracy. Decision-curve analysis was used to compare the net benefit of U_clot-guided triage with “treat-all” and “treat-none” strategies.^41^ Statistical analyses were performed in Python and/or R.

## Results

### Cohort Characteristics and Training Setup

We trained a 3D full-resolution nnU-Net using 5-fold cross-validation, with each fold optimized for 1200 epochs organized into three equal learning-rate cycles of 400 epochs. The overall training cohort comprised 223 MT-treated AIS patients with baseline NCCT and CTA. Across these training cases, ground-truth clot volume was 78.7 ± 62.6 mm³ (mean ± SD), and the median HU within the clot mask was 56.0 ± 7.7 HU. Using our predefined categories (small: 0–<50 mm³, medium: 50–<150 mm³, large: ≥150 mm³; hyperdense: median HU > 51), the training set included 91 small, 108 medium, and 24 large clots, of which 170 were hyperdense and 53 non-hyperdense.

The held-out test cohort consisted of 80 patients. In this set, the average ground-truth clot volume was 81.7 ± 61.7 mm³, with median clot density of 56.9 ± 7.8 HU, closely matching the training distribution. By size, the test cohort contained 33 small, 36 medium, and 11 large clots; by density, 65 thrombi were hyperdense and 15 non-hyperdense. Representative examples of clot appearance and automated segmentations across these phenotypes are shown in Figure 1.

**Figure 1.**
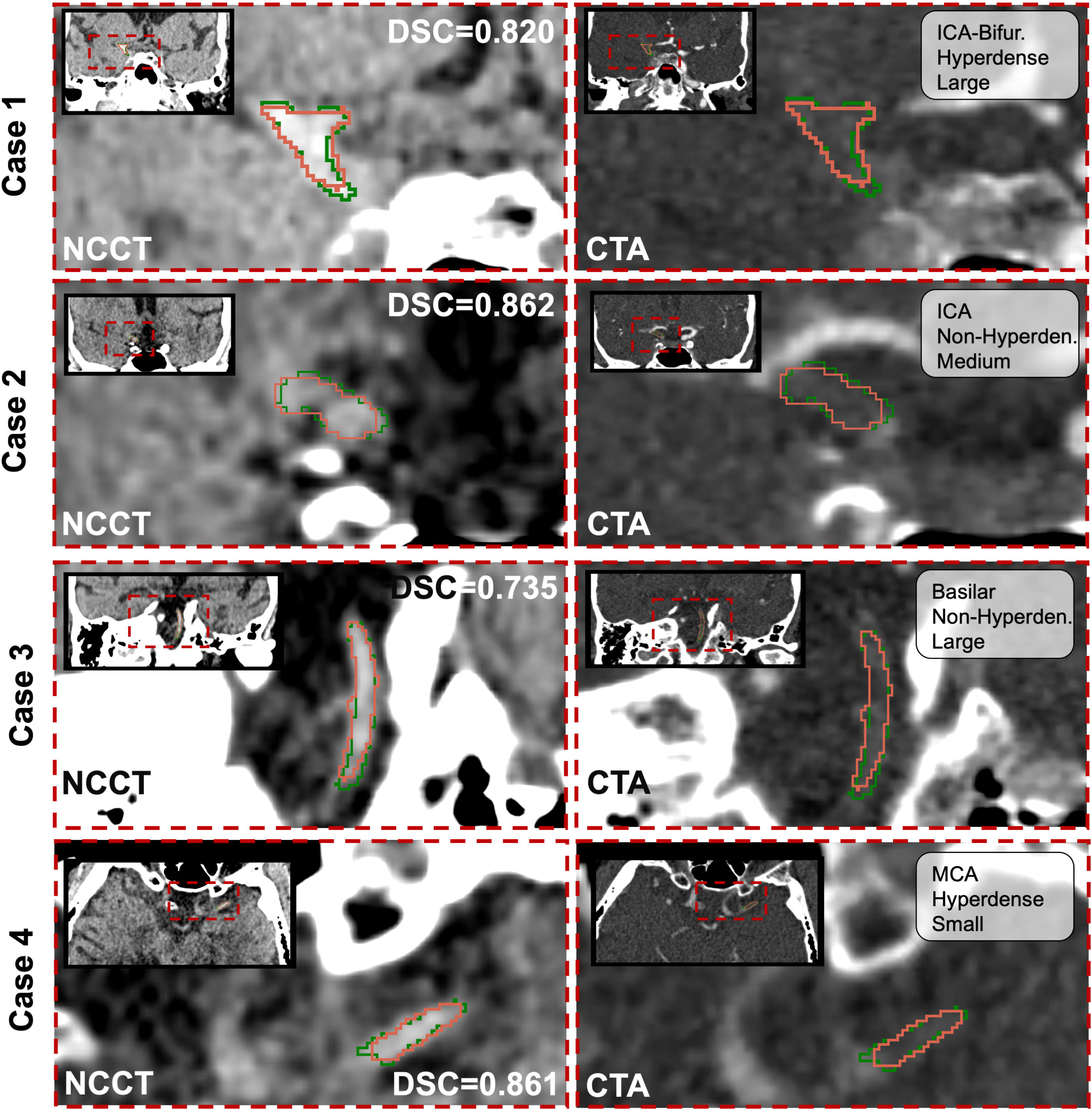
Representative examples of intracranial thrombus appearance and segmentation across clot phenotypes. **Left column:** NCCT images for four example cases (rows: Case 1–4), each shown as a zoomed-in view of the clot region with a smaller zoomed-out inset in the corner for anatomical context. Manual ground-truth clot segmentations are shown in green, and automated model predictions in red. For each case, the Dice similarity coefficient (DSC) between the automated and manual clot segmentation is indicated on the NCCT panel. **Right column:** Corresponding CTA images for the same four cases, again shown with zoomed-in clot views and zoomed-out insets. CTA panels are annotated with clot location (e.g., MCA, ICA, basilar), hyperdense versus non-hyperdense status, and volume category (small, medium, or large). Cases 1–3 are displayed in the coronal plane, and Case 4 is displayed in the axial plane, illustrating the diversity of clot size, density, location, and segmentation performance captured by the model. *Abbreviations: CTA = computed tomography angiography; DSC = dice similarity coefficient; NCCT = non-contrast computed tomography; ICA = internal carotid artery; ICA-Bifur. = internal carotid artery bifurcation; MCA = middle cerebral artery*.

### Segmentation Performance

#### Overall Segmentation Performance

Across the full test cohort (n = 80), the model successfully detected clot regions (≥1 voxel overlap with ground truth) in 92.5% of cases (74/80). False positives (model predicted clot but no overlap with ground truth) occurred in 3.75% (3/80), and false negatives (no model prediction despite ground-truth clot) in 3.75% (3/80). A representative false-negative example (small non-hyperdense M2 clot with no predicted segmentation) is shown in Supplemental Figure 5. For the cohort, the model predictions resulted in an average Dice coefficient of 0.638± 0.242, an average surface Dice at 1 mm of 0.646 ± 0.277, and an IoU of 0.507 ± 0.223.

Precision and recall were 0.646± 0.277and 0.696 ± 0.259, respectively, and volumetric similarity was 0.779 ± 0.235. Overall, 70% of cases (56 / 80) achieved a Dice ≥ 0.60 and 57.5% (46 / 80) achieved a Dice ≥ 0.70.

#### Volumetric Agreement Between Ground-Truth and Predictions

Predicted clot volumes showed good agreement with ground-truth volumes (Figure 3A and Supplemental Figure 1A). The average ground-truth volume was 81.663 ± 61.701 mm^3^, and the average predicted clot volume was 86.927 ± 62.012 mm^3^. Bland-Altman analysis demonstrated a mean value difference ΔV = 5.5 mm^3^ (95% limits of agreement, -64.8 to 75.8 mm^3^), with no strong systematic bias over the range of clot sizes (Figure 3A). Predicted and ground-truth volumes correlated strongly (Spearman *ρ* = 0.772, p < 0.001; Supplemental Figure 1A). Linear regression yielded a slope of β = 0.84 and intercept of α = 18.7, with an R² of 0.693, indicating good calibration. The intraclass correlation coefficient (ICC, two-way mixed, absolute agreement) for volumes was 0.831, indicating good agreement.

**Figure 2.**
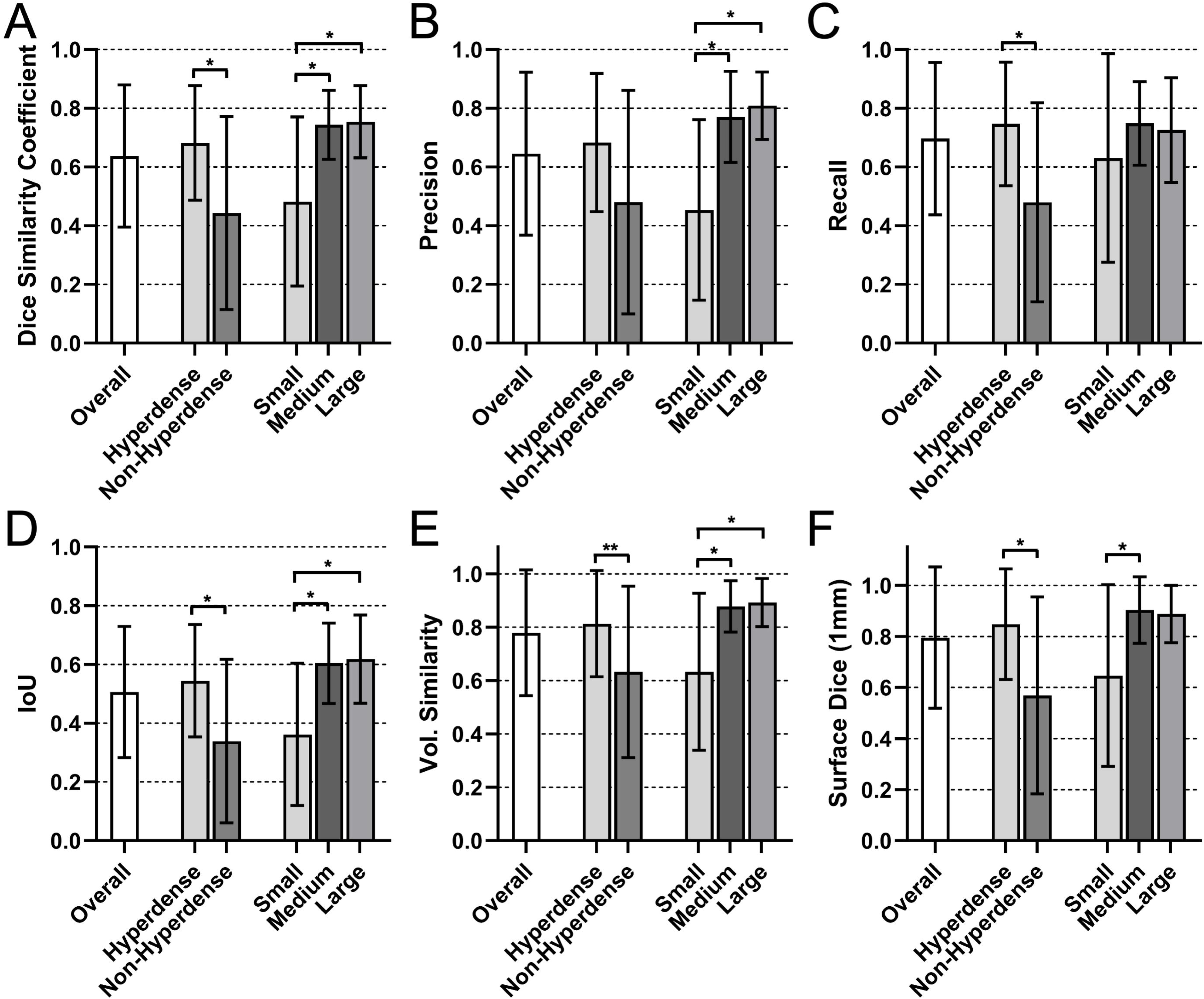
Segmentation performance across clot density and size subgroups. Panels show distributions of segmentation metrics for the overall test cohort, stratified by thrombus density (hyperdense vs non-hyperdense) and by clot size (small, medium, large). **(A)** Dice similarity coefficient. **(B)** Precision. **(C)** Recall (sensitivity). **(D)** Intersection over Union (IoU). **(E)** Volumetric similarity. **(F)** Surface Dice at 1 mm. Each panel summarizes the metric for the full test set and then compares hyperdense vs non-hyperdense thrombi, as well as small (0–<50 mm³), medium (50–<150 mm³), and large (≥150 mm³) clots, highlighting systematic differences in segmentation performance across clot phenotypes. *Abbreviations: IoU = intersection over union; Vol. = volume; mm = millimeters*.

**Figure 3.**
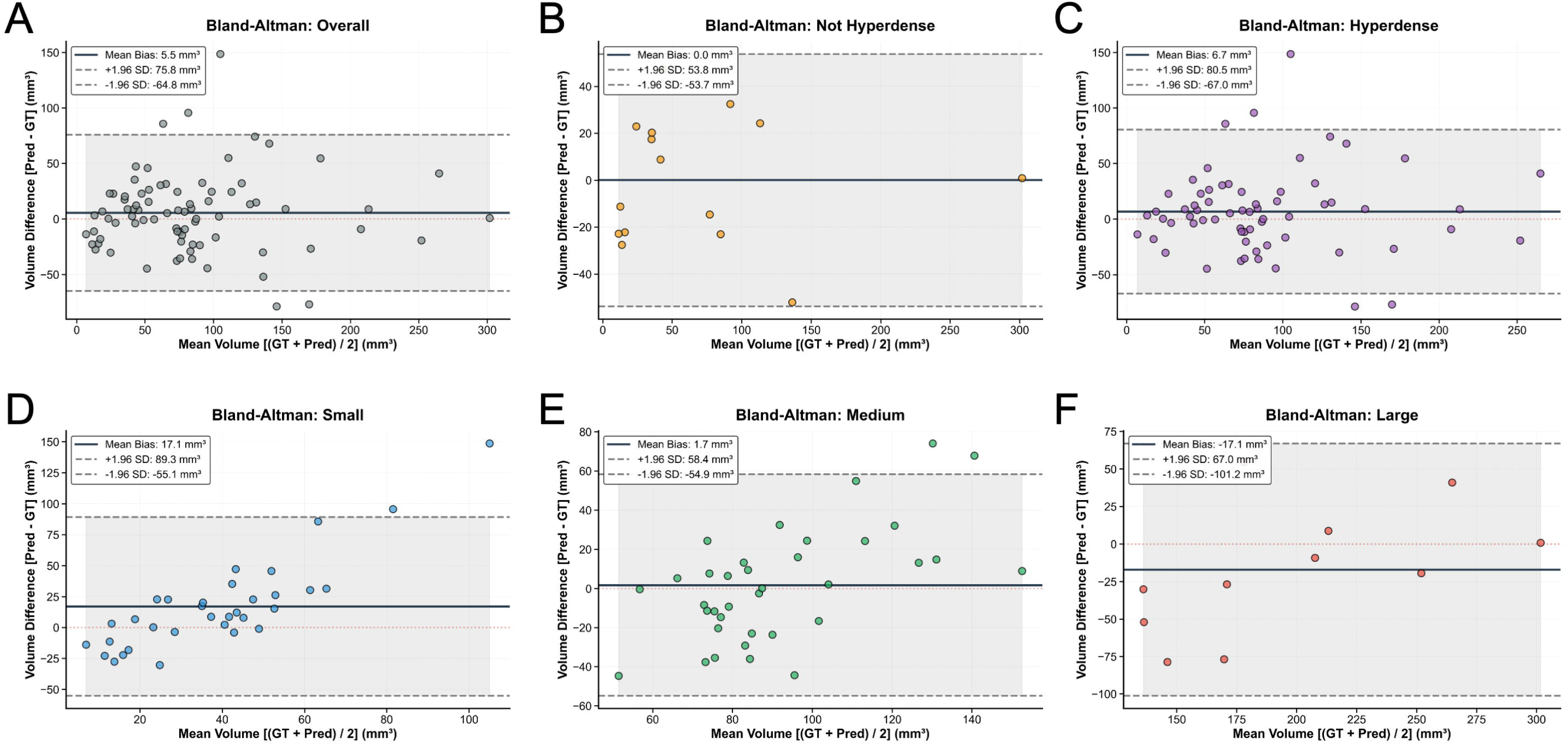
Bland–Altman analysis of clot volume agreement between ground-truth and predicted segmentations across cohorts. **(A)** Overall test cohort. **(B)** Non-hyperdense clots. **(C)** Hyperdense clots (defined as median NCCT HU ≥ 51). **(D)** Small clots (0–<50 mm³). **(E)** Medium clots (50–<150 mm³). **(F)** Large clots (≥150 mm³). For each panel, the x-axis shows the mean of ground-truth (GT) and predicted (Pred) clot volumes, and the y-axis shows the volume difference (Pred – GT). The solid horizontal line indicates the mean bias, and the dashed lines indicate the ±1.96 SD limits of agreement. *Abbreviations: GT = ground truth; Pred = predicted; SD = standard deviation; mm³ = cubic millimeters; HU = Hounsfield units*.

#### Performance Across Clot Density and Size

When stratified by thrombus density on NCCT, hyperdense clots (median HU > 51; n = 65) had higher segmentation quality than non-hyperdense clots (n = 15). Average Dice was 0.683 ± 0.194 vs 0.443 ± 0.329 (p = 0.005), surface Dice at 1 mm 0.855 ± 0.216 vs 0.579 ± 0.391 (p = 0.003), and volumetric similarity 0.813 ± 0.199 vs 0.633 ± 0.321 (p = 0.015) for hyperdense vs non-hyperdense thrombi, respectively (Figure 2). Segmentation performance also varied with clot size (Figure 2). For small clots (0 < V < 50 mm³; n = 33), medium clots (50 ≤ V < 150 mm³; n = 36), and large clots (V ≥ 150 mm³; n = 11), average Dice was 0.483 ± 0.288, 0.744 ± 0.117, and 0.754 ± 0.123, respectively (overall p < 0.001). Pairwise comparisons showed significantly lower Dice for small clots vs medium (p<0.001) and large clots (p=0.004); no significant difference between medium vs large clots (p=0.999). Similar patterns were observed for surface Dice, recall, and volumetric similarity (Figure 2).

Volumetric agreement, assessed by Bland–Altman demonstrated a mean value difference for non-hyperdense and hyperdense clot cohorts of 0 mm^3^ (95% limits of agreement, - 53.7 to 53.8 mm^3^) and 6.7mm^3^ (95% limits of agreement, -67.0 to 80.5mm^3^) respectively, as shown in Figure 3B-C. Mean value differences from Bland-Altman analyses for small, medium and large clots were 18.1 mm^3^ (95% limits of agreement, -55.1 to 89.3 mm^3^), 1.7 mm^3^ (95% limits of agreement, -54.9 to 58.4 mm^3^), and -17.1 mm^3^ (95% limits of agreement, -101.2 to 67.0 mm^3^) respectively, shown in Figure 3D-F, suggesting that the volumes of smaller clots were generally over-predicted whereas for the larger clots, were under-predicted. The ICC calculated using a two-way mixed effect, absolute-agreement model was 0.831 (good) for the overall cohort. Across cohorts stratified based on hyperdensity, the ICCs for non-hyperdense clots were 0.940 (excellent) and 0.788 (good) for non-hyperdense clots. Subgroup-specific volume correlations and regression lines are shown in Supplemental Figure 1B–F. Across clot sized based on volumes, small clots showed poorest agreement with an ICC of 0.074 (poor), followed by 0.458 (poor) for medium clots and 0.724 (moderate) for large clots.

### Uncertainty and Segmentation Quality

#### Overall Relationship Between Uncertainty and Segmentation Performance

For the cases where the model predicted some clot ROI (n=77/80, i.e. 96.25% of the cases), the clot-level uncertainty score (U_clot) was inversely associated with segmentation quality. The correlation between U_clot and Dice was *ρ* = –0.704 (Spearman, p < 0.001; Figure 4B). The agreement between U_clot and Dice was −0.820 (negative indicating inverse association), as quantified via ICC. Qualitative examples of voxel-wise entropy maps highlighting regions of disagreement between predicted and ground-truth masks, and elevated uncertainty along clot boundaries, are shown in Figure 4A.

**Figure 4.**
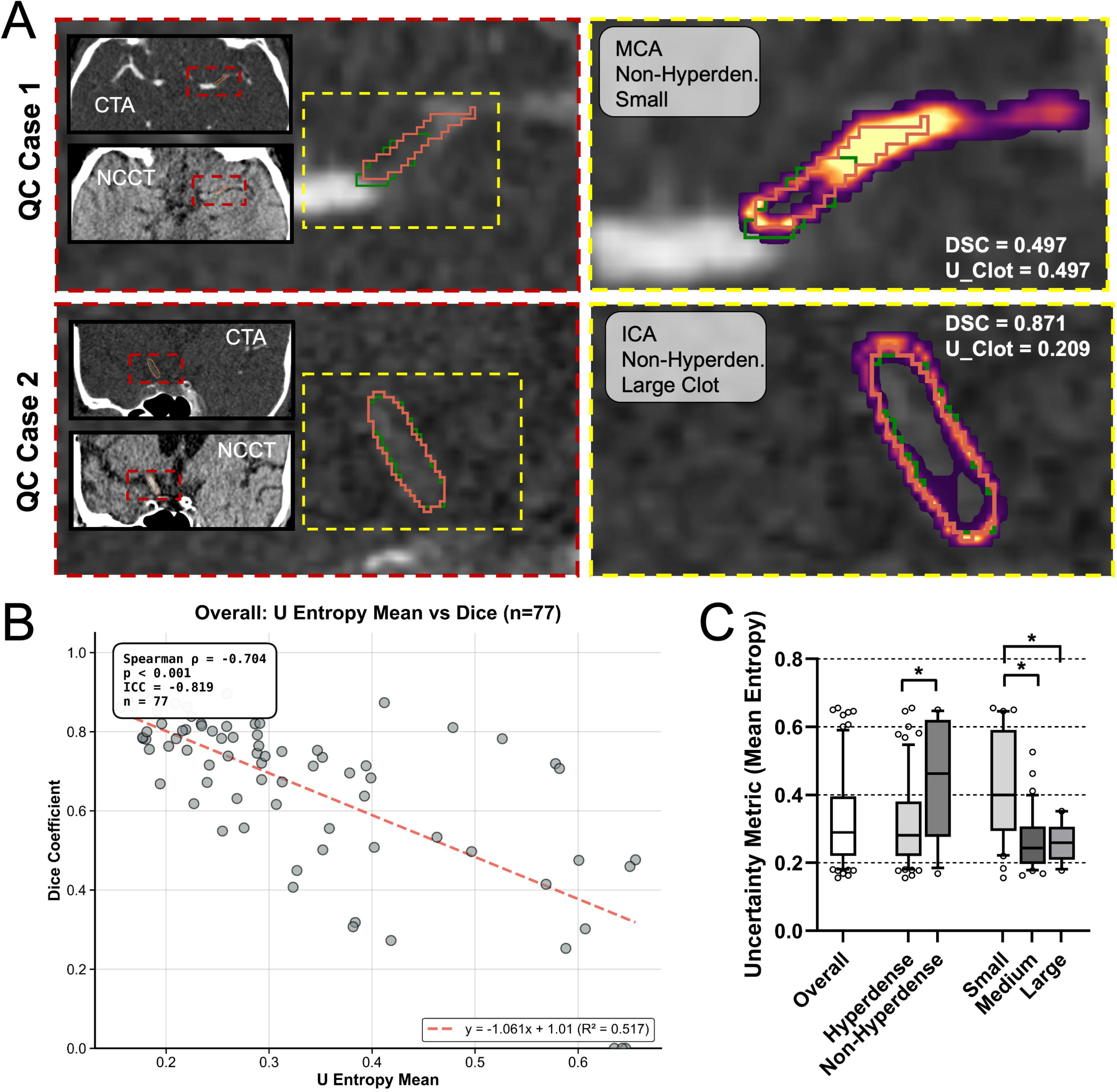
Uncertainty behavior and its relationship to clot segmentation quality. **(A)** Representative quality-control (QC) examples illustrating voxel-wise entropy. For each case, the left panel shows the CTA with ground-truth (GT, green) and predicted (Pred, red) clot masks overlaid, with a zoomed-in view of the thrombus and inset NCCT/CTA context images. The right panel shows the same CTA slice with the entropy map overlaid and GT/Pred contours. In QC case 1, entropy peaks align with regions of mismatch between GT and Pred. In QC case 2, entropy is highest along the clot boundary, where contour uncertainty is expected. The Dice similarity coefficient (DSC) and clot-level uncertainty score (U_clot) are indicated for each case. **(B)** Scatter plot of DSC versus U_clot for all test cases with a non-empty prediction (n = 77), with a fitted linear regression line (dashed red). The Spearman correlation coefficient (ρ), corresponding p value, and intraclass correlation coefficient (ICC) summarize the strong inverse association between Dice and U_clot. **(C)** Distribution of U_clot across the overall test cohort and stratified by clot phenotype: hyperdense versus non-hyperdense thrombi and small, medium, and large clots based on predefined volume thresholds, illustrating systematically higher uncertainty in non-hyperdense and smaller clots. *Abbreviations: QC = quality control; CTA = computed tomography angiography; NCCT = non-contrast computed tomography; Non-Hyperden. = non-hyperdense; MCA = middle cerebral artery; ICA = internal carotid artery; DSC = dice similarity coefficient; U_clot = clot-level uncertainty score; ICC = intraclass correlation coefficient; n = number of cases*.

#### Uncertainty Across Clot Density and Size Categories

Hyperdense thrombi tended to have lower uncertainty than non-hyperdense thrombi (0.310 ± 0.126 [n=64] vs 0.439 ± 0.177 [n=13], p = 0.015; Figure 4C), mirroring the higher Dice observed in hyperdense clots. Similarly, U_clot differed significantly across clot size categories (overall p < 0.001; Figure 4C). Small clots showed the highest uncertainty (0.433 ± 0.160, n=30), followed by medium (0.268 ± 0.088, n=36) and large (0.262 ± 0.059, n=11) clots; pairwise comparisons showed small clots had significantly higher uncertainty compared to medium (p < 0.001) and large clots (p = 0.008), whereas there were no significant differences between uncertainty values from large and medium clots.

Within each subgroup, U_clot remained a meaningful indicator of segmentation quality. For hyperdense clots, the correlation between U_clot and Dice was *ρ* = –0.605 (Spearman, p < 0.001), and for non-hyperdense clots r = –0.905 (Pearson, p < 0.001). Similarly, for small, medium, and large clots, correlations between U_clot and Dice were *ρ* = –0.740 (Spearman, p < 0.001), *ρ* = –0.515 (Spearman, p < 0.001), and r = –0.669 (Pearson, p = 0.024), respectively, indicating that higher uncertainty consistently marked poorer segmentations regardless of clot phenotype, as illustrated in Supplemental Figure 2. The inverse agreements (quantified via ICC), also ranged from −0.607 to −0.936 (See Supplemental Figure 2), suggesting good to excellent agreements between the U_clot metric and performance of segmentation quantified by Dice.

### Uncertainty-Based Triage

#### Optimal Uncertainty Cut-off for Identifying Poorly Segmented Cases

Using a Dice threshold of 0.60 to define “well segmented” (Dice ≥ 0.60) versus “poorly segmented” (Dice < 0.60) cases, U_clot showed good discriminative ability (Figure 5A). The ROC curve for U_clot as a univariate predictor yielded an AUC of 0.892. The optimal U_clot cut-off derived from Youden’s index was U_clot = 0.323. At this threshold, the confusion matrix for predicting poor segmentations showed a sensitivity of 90.476% (19/21), specificity of 78.571% (44/56), and an overall accuracy of 81.818% (63/77). Positive and negative predictive values were 61.290% (19/31) and 95.652% (44/46), respectively (Figure 5A and Supplemental Figure 3A). When stratified by clot phenotype, U_clot maintained excellent discriminative performance, with AUCs of 0.844 and 1.000 for hyperdense and non-hyperdense clots, respectively, and 0.857, 0.828, and 1.000 for small, medium, and large clots (Figure 5B-C). Across all stratifications, the algorithm showed a strong ability to identify cases requiring manual review.

**Figure 5.**
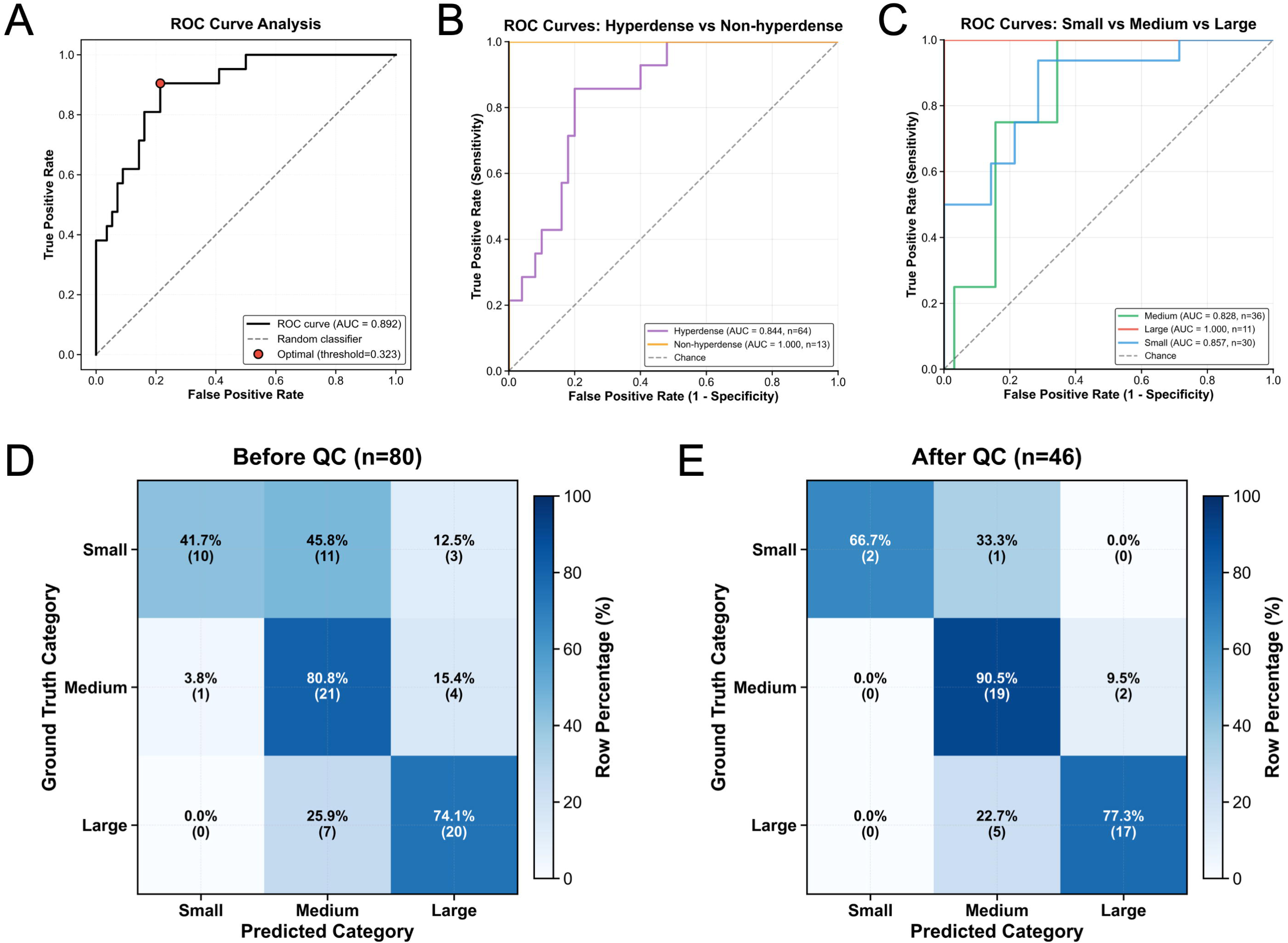
Uncertainty-based triage and impact on volume-based size categorization. **(A)** Receiver operating characteristic (ROC) curve for using the clot-level uncertainty score (U_clot) to distinguish well-segmented (Dice ≥ 0.60) from poorly segmented clots, with area under the curve (AUC) and the optimal threshold (Youden’s index) indicated. **(B–C)** ROC curves of U_clot stratified by clot phenotype: **(B)** hyperdense versus non-hyperdense thrombi, and **(C)** small, medium, and large clots (volume-based categories). Corresponding AUCs are shown in the legends, demonstrating robust discrimination across subgroups. **(D–E)** Three-by-three confusion matrices for predicted versus ground-truth volume-based size categories (small/medium/large): **(D)** full test cohort, accuracy 0.662 and Cohen’s κ = 0.490; **(E)** restricted to cases passing uncertainty-based QC (U_clot below the optimal threshold), with accuracy 0.826 and Cohen’s κ = 0.685, indicating improved categorical agreement after triage. *Abbreviations: ROC = receiver operating characteristic; AUC = area under the curve; QC = quality control*.

Because our intended use is to safely triage segmentations for expert inspection, we focused on sensitivity (ability to catch poor segmentations) and negative predictive value (NPV) (reassurance that automatically accepted cases are truly adequate). Within the hyperdense cohort, the confusion matrix for predicting poor segmentation yielded a sensitivity of 85.714% (12/14) and an NPV of 95.238% (40/42). For non-hyperdense clots, both sensitivity and NPV were 100% (7/7 and 4/4, respectively).

Similarly, for small clots, sensitivity was 93.75% (15/16) with an NPV of 88.889% (8/9). For medium clots, sensitivity and NPV were 75% and 96.429%, respectively, and for large clots both sensitivity and NPV reached 100% (Supplemental Figure 3). Taken together, these results indicate that, even within challenging sub-cohorts, U_clot reliably flags most poor segmentations while maintaining a high probability that unflagged cases are of acceptable quality.

#### Trade-off between Automation and Quality

Varying the U_clot threshold defined different operating points on the automation–quality trade-off (Figure 6). These curves were plotted by sampling 20 equidistant U_Clot values, with the minimum and maximum for U_Clot threshold set to wherever in the low or high uncertainty cohorts there’s at least 5 cases for obvious reasons. Under these criteria, as the threshold for U_clot decreased (more stringent triage), the proportion of cases automatically accepted fell from 94.805% to 5.195%, while the mean Dice among accepted cases increased from 0.686 to 0.848.

**Figure 6.**
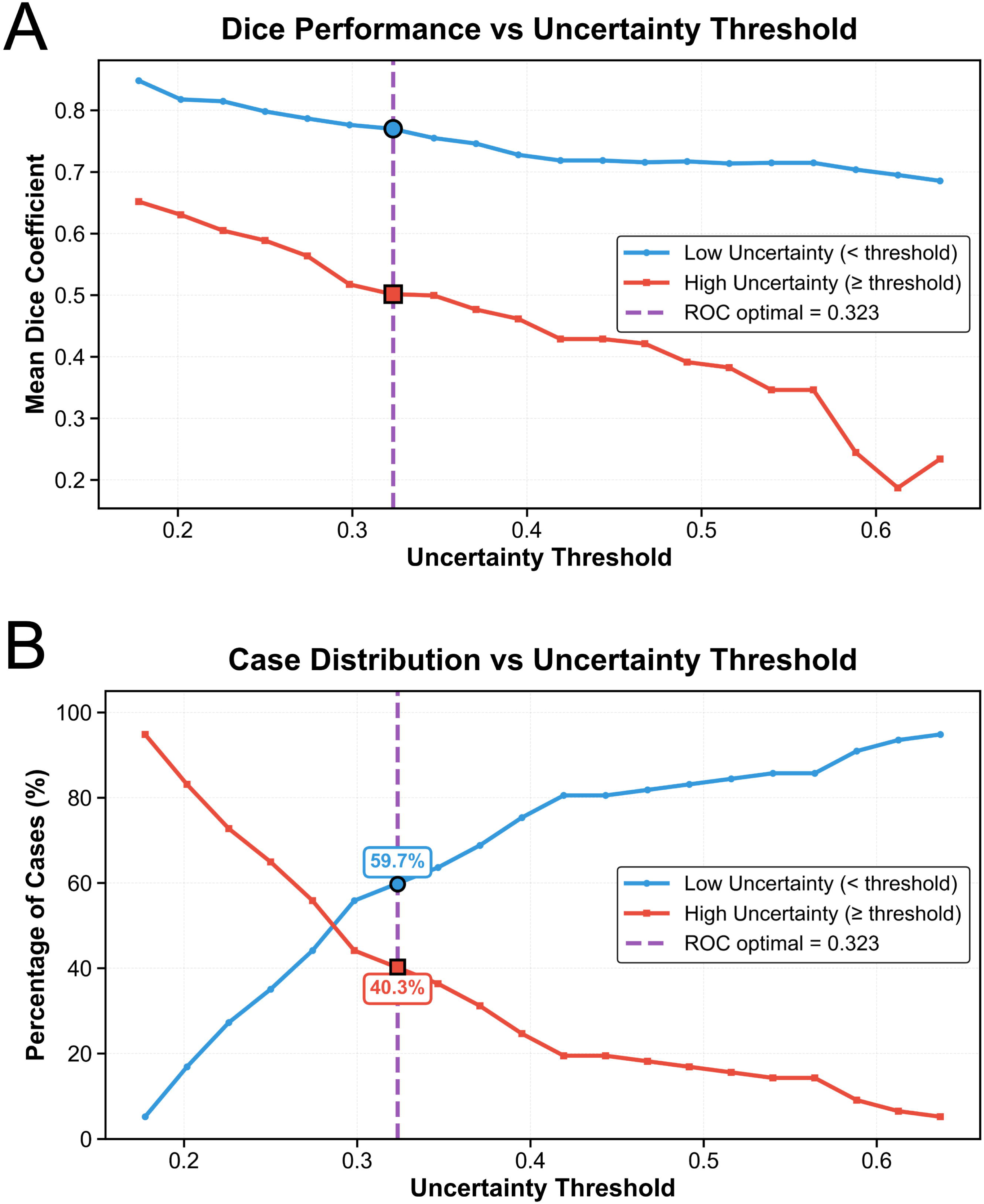
Automation–quality trade-off as a function of the uncertainty threshold. **(A)** Mean Dice similarity coefficient (DSC) of accepted cases as a function of the U_clot threshold. As the threshold is lowered (stricter triage), mean DSC among included (low-uncertainty) cases increases. The operating point at U_clot = 0.323 is highlighted. **(B)** Proportion of cases included (low-uncertainty) versus triaged (high-uncertainty) across the same range of U_clot thresholds. At U_clot = 0.323, approximately 60% of cases are retained for fully automated use, while ∼40% are flagged for manual review. *Abbreviations: DSC = Dice similarity coefficient; U_clot = clot-level uncertainty score*.

At the optimal threshold U_clot = 0.323, 59.740% (46/77) of cases would have been accepted without expert review and 40.260% (31/77) flagged for manual inspection (Figure 6). Among accepted cases, average Dice was 0.771 ± 0.085, compared with 0.502 ± 0.238 among triaged cases (p < 0.001). Similar improvements were observed for surface Dice (0.933 ± 0.085 vs 0.689 ± 0.295, p < 0.001) and volumetric similarity (0.889 ± 0.096 vs 0.693 ± 0.212, p < 0.001).

#### Decision-curve analysis for uncertainty-based triage

We evaluated the clinical utility of U_clot as a triage rule using decision-curve analysis (Supplemental Figure 4). The strategies of reviewing no segmentations (“treat none as high-risk”) and reviewing all segmentations (“treat all as high-risk”) both yielded a net benefit of approximately zero across the entire range of uncertainty thresholds, indicating no advantage over each other in this setting.

By contrast, the uncertainty-based stratification strategy (blue curve) provided positive net benefit over a wide range of thresholds. Net benefit increased as the uncertainty threshold rose from ∼0.18 and reached a maximum at the ROC-derived optimal cut-off U_clot = 0.323, where the net benefit was 0.188. This corresponds to an effective gain of about 19 additional poorly segmented cases correctly flagged for manual review per 100 patients, compared with either reviewing all cases or reviewing none, after accounting for unnecessary reviews of adequately segmented cases.

Beyond this point, net benefit gradually declined as thresholds increased (reflecting more missed poor segmentations), but remained above the “treat all/none” strategies over most of the clinically relevant threshold range. These findings support the chosen threshold of U_clot = 0.323 as a clinically efficient operating point for uncertainty-based triage.

### Effect of Triage on Categorical Clot Size Classification

#### Agreement in Volume-based Size Categories

Using ground-truth and predicted volumes discretized into small (0–50 mm³), medium (50–150 mm³), and large (≥150 mm³) categories, the model correctly matched the ground-truth size category in 66.234% of cases (51/77; 3×3 confusion matrix, Figure 5D). Overall, Cohen’s κ was 0.490, indicating moderate agreement. Most misclassifications occurred between adjacent categories (small vs medium or medium vs large), while “jumping” errors from small directly to large (or vice versa) were relatively rare, occurring in 3.896% of cases (3/77; Figure 5D).

#### Improvement with Uncertainty-based Triage

When the analysis was restricted to cases with U_clot below the optimal triage threshold, categorical size agreement improved substantially. In this “low-uncertainty” subset (n = 46/77), accuracy increased to 82.609%, and Cohen’s κ rose to 0.685 (Figure 5E), consistent with substantial agreement between predicted and ground-truth volume categories. Notably, all three small-to-large misclassifications (3.9% of cases) were successfully triaged out, demonstrating that uncertainty-based filtering eliminates the most clinically problematic volume estimation errors. Together, these findings indicate that uncertainty-based triage can meaningfully enhance the reliability of automated clot size categorization, by preferentially retaining cases with both accurate segmentations and stable volume estimates.

## Discussion

In this study, we developed and evaluated an uncertainty-aware deep learning framework for automated intracranial thrombus segmentation using NCCT and CTA in stroke patients. Our nnU-Net–based model achieved an average Dice of 0.64 with good volumetric agreement (ICC 0.83) between predicted and manual clot volumes. Segmentation quality varied by clot phenotype, with higher average performance for hyperdense (median ROI HU > 51) and medium-to-large thrombi (>50 mm³) and lower performance for small or non-hyperdense clots. Using an ensemble-based entropy measure summarized as a clot-level uncertainty score (U_clot), we showed that uncertainty was tightly and consistently linked to segmentation performance across phenotypes, and that a single cut-off on U_clot could be used to triage segmentations for manual review with high sensitivity and negative predictive value for the first time in intracranial thrombus segmentation. When applied as a triage rule, U_clot allowed roughly 60% of cases to be processed fully automatically while substantially improving both volumetric agreement and agreement in clinically relevant volume-based size categories.

The segmentation performance measured here is in line with and extends previous work on fully automated thrombus segmentation from NCCT and CTA. Mojtahedi et al. proposed a dual-encoder 3D U-Net with NCCT and CTA as separate input streams, combined with a dynamic bounding-box strategy; in a 100-patient test set they reported a Dice of 0.62, surface Dice of 0.78, and an ICC of 0.60 for thrombus volume, with 4% of cases missed entirely.^17^ Zhu et al. used a coarse-to-fine deep learning approach to segment thrombus and occlusion location on CTA, obtaining Dice scores around 0.71 in their test cohort. Our mean Dice of 0.64 therefore falls squarely within the range of these prior reports, despite differences in dataset size, scanner mix, and model design.^16^ Notably, in our cohort the volumetric ICC for the overall group (0.83) was numerically higher than that reported by Mojtahedi et al. (0.60), suggesting that our predicted clot volumes showed closer agreement with expert measurements despite similar Dice scores and despite our more diverse cohort (including smaller/non-hyperdense clots where performance is inherently challenging).^17^

We observed that clot size and CT density were important determinants of segmentation performance. This is consistent with previous work: Mojtahedi et al. reported that hyperdense clots and larger volumes were more easily segmented and that performance degraded in low-contrast or very small thrombi, and in our cohort Dice, surface Dice, recall, volumetric similarity, and volumetric ICC were all higher for hyperdense than for non-hyperdense clots and for medium/large than for small clots. Our Bland–Altman analyses further showed a tendency to overestimate volumes in small clots and underestimate large clots, indicating that segmentation error is not uniform across the volume spectrum. At the same time, several studies, including our own prior work, have shown that clot annotations themselves exhibit only modest agreement between observers: Mojtahedi et al. reported a mean pairwise Dice of 0.47 (95% CI 0.33–0.61) across four readers, Zhu et al. found Dice values as low as ∼0.54–0.58 between individual raters and a STAPLE consensus, and our earlier inter-user analysis yielded pairwise Dice of ∼0.56–0.60 with average surface distances on the order of 1–2 voxels.^16–18^ Together, these findings indicate that automated thrombus segmentation performs best for larger, hyperdense clots and that both human and model performance are influenced by how clearly clot boundaries are depicted on CT.

Given these challenges, and the fact that human observers themselves only achieve modest agreement when segmenting clots, it is not sufficient to report a single average Dice score for an automated model. In practice, we also need to know, case by case, how confident we can be in the segmentation that the network produces. To address this, we focused on

uncertainty quantification. Our framework uses the ensemble-derived U_clot score as a case-level confidence measure that reflects how stable the predicted clot mask is across posterior samples. Instead of treating every predicted mask as equally reliable, U_clot allows us to identify cases in which the segmentation is likely unstable and at higher risk of misleading downstream volume-based decisions, while allowing high-confidence cases, where humans and the model are more likely to agree, to be processed automatically. Prior automated thrombus segmentation studies have treated the model output as deterministic and have not attempted to quantify, for each patient, when an individual segmentation may be unreliable. For example, Mojtahedi et al. used their automated pipeline to derive thrombus volume, density, perviousness, and heterogeneity in large multicenter cohorts and showed meaningful associations with clinical and technical outcomes, but their analyses implicitly assumed adequate segmentation quality for all cases.^42^ Our work complements these efforts by explicitly characterizing how segmentation error varies across clot phenotypes and introducing a practical, data-driven uncertainty metric that can be used to gate the use of automated thrombus measurements.

A key contribution is demonstrating that U_clot, an ensemble-based entropy metric, provides a robust, clot-phenotype-agnostic indicator of segmentation quality across the challenging spectrum of clinical thrombi. Across the full cohort, U_clot was strongly and inversely correlated with Dice, and this relationship held within hyperdense and non-hyperdense subgroups and across small, medium, and large clots. High uncertainty reliably marked cases with low Dice, low volumetric similarity, and poor surface agreement, whereas low-uncertainty cases clustered at higher segmentation quality. This alignment is crucial: if uncertainty did not decrease as segmentation quality improved, it would not be useful as a quality-control signal.

Instead, our data show that U_clot encodes meaningful information about segmentation reliability rather than random model variability, consistent with prior work on trajectory-based posterior approximation and uncertainty-aware nnU-Net variants in other domains. From a clinical perspective, this translated into a usable triage rule: using Dice ≥ 0.60 as an “acceptable” segmentation, U_clot achieved an AUC of 0.89, and at the chosen threshold of 0.323 it provided high sensitivity (∼90%) and a high negative predictive value (∼96%) for detecting poor segmentations. Applying this threshold, approximately 60% of cases could be accepted without manual review, and agreement in volume-based size categories improved from about 66% (κ = 0.49) to 83% (κ = 0.69), with extreme misclassifications from small directly to large clots disappearing. Together, these findings suggest that uncertainty estimates can both flag most problematic segmentations and materially improve the reliability of automated clot measurements without requiring manual inspection in every patient.

This study has several limitations. It was conducted at a single center with modest sample size and relatively uniform imaging protocols dominated by one scanner platform, so generalizability to broader multicenter cohorts remains to be established.^43^ Ground-truth masks were generated by one primary rater under supervision rather than by multi-rater consensus; although prior work from our group has shown reasonable inter-observer agreement for clot segmentation, residual label noise may have affected both training and evaluation.^31,44^ The U_clot threshold was derived post hoc using ROC analysis in this dataset, which may overestimate performance, and therefore requires confirmation in an independent cohort and, ideally, prospective testing in real-world workflows. Methodologically, we evaluated a single 3D nnU-Net configuration and one ensemble-based entropy measure; other approaches such as test-time augmentation, independently trained deep ensembles, or more explicit decompositions of aleatoric and epistemic uncertainty could provide complementary information and warrant comparison.^45–48^ We also restricted the ROI to the major intracranial arteries and examined uncertainty mainly in relation to segmentation quality and volume-based size categories. Future work should extend these analyses to more distal occlusions and posterior circulation strokes and assess how U_clot and related measures influence the robustness and clinical utility of thrombus radiomics, perviousness, and outcome prediction models.

## Conclusions

In summary, we present an uncertainty-aware deep learning framework for automatic thrombus segmentation on NCCT and CTA that achieves human-level performance while providing a robust, interpretable per-case measure of confidence. By leveraging this uncertainty metric to triage segmentations for manual review, we can substantially improve volumetric agreement and size-category classification, particularly for challenging clot phenotypes, while still enabling automation for most patients. These results suggest that explicit uncertainty modeling and triage may be a viable path toward safe deployment of automated clot segmentation in both clinical practice and large-scale stroke research. More broadly, embedding such uncertainty-aware quality control into segmentation pipelines addresses a critical gap in the workflow of future AI imaging tools for stroke care, where reliable, case-specific confidence estimates will be essential for clinical adoption.

## Data Availability

All data produced in the present study are available upon reasonable request to the authors.

## Acknowledgements

Funding for this work was provided in part by the Society of Vascular and Interventional Neurology’s Pilot Research Grant Program. Additionally, part of the computational resources was provided by the Center for Computational Research at the University at Buffalo.^49^

