## Supplemental Figures for "Automated Intracranial Thrombus Segmentation from CT Images of Patients with Acute Ischemic Stroke: A Dual-Channel nnU-Net Approach with Uncertainty Quantification"

[a] Department of Neurosurgery, University at Buffalo, NY, USA; [b] Department of Pathology and Anatomical Sciences, University at Buffalo, Buffalo, NY, USA; [c] Canon Stroke and Vascular Research Center, Buffalo, NY, USA; [d] Department of Interventional Neuroradiology, University of California, Los Angeles, CA, USA; [e] Department of Neurosurgery and Brain Repair, University of South Florida, Tampa, FL, USA; [f] Department of Neurology, University of Texas Health, Houston, TX, USA

***Corresponding Author:**

Tatsat R. Patel, PhD
875 Ellicott Street
Canon Stroke and Vascular Research Center University at Buffalo
Buffalo, NY 14203, USA


**Running Title:** Uncertainty-Aware CT Thrombus Segmentation

**Keywords:** Acute ischemic stroke, Intracranial thrombus, CT angiography, Deep learning, Uncertainty quantification

### Supplemental Figures

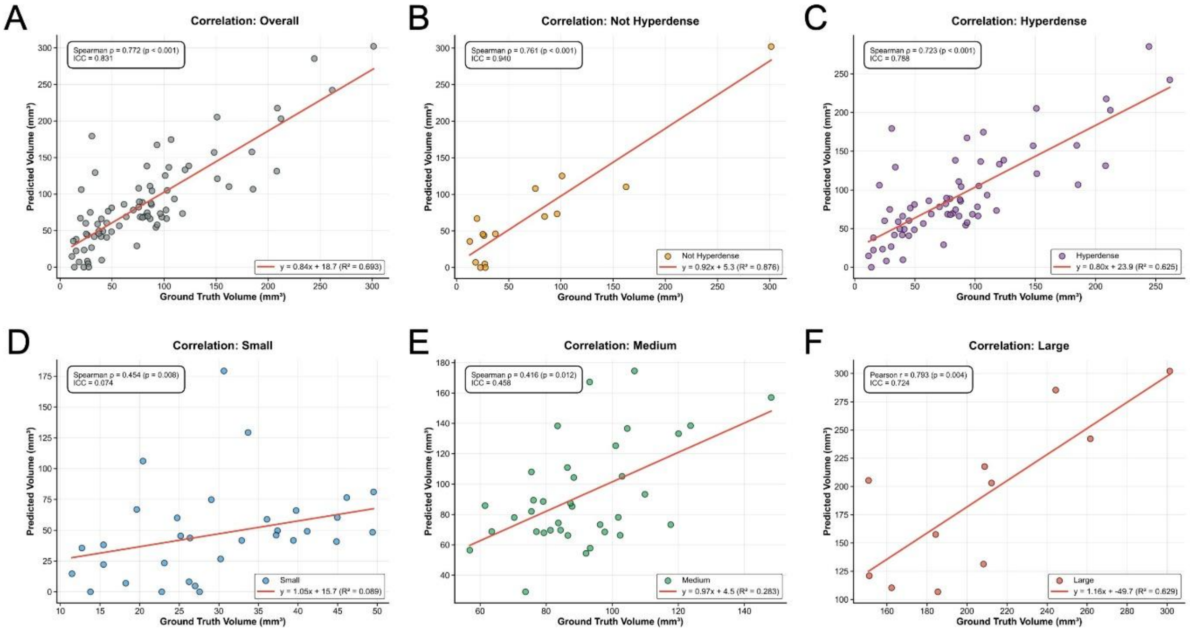


**Supplemental Figure 1. Correlation and calibration between predicted and ground-truth clot volumes.** Scatterplots of predicted versus ground-truth clot volume for **(A)** the overall test cohort, **(B)** non-hyperdense clots, **(C)** hyperdense clots (median NCCT HU ≥ 51), and by size category: **(D)** small (0–<50 mm³), **(E)** medium (50–<150 mm³), and **(F)** large (≥150 mm³) clots. Each panel shows the fitted linear regression line (red), regression equation, and coefficient of determination ($R^2$), along with the corresponding Pearson or Spearman correlation coefficient (chosen based on normality), p value, and intraclass correlation coefficient (ICC) for volumetric agreement. *Abbreviations: ICC = intraclass correlation coefficient; mm³ = cubic millimeters.*

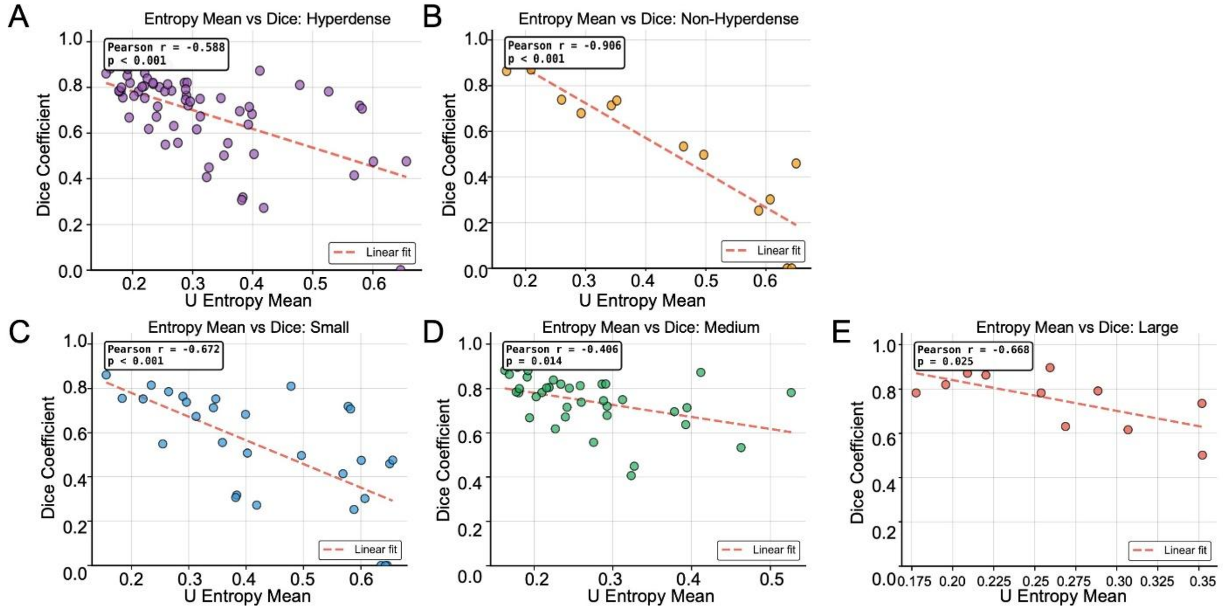


**Supplemental Figure 2. Relationship between clot-level uncertainty and segmentation quality within clot subgroups.** Scatterplots of Dice similarity coefficient (DSC) versus case-level clot uncertainty (U_clot) for **(A)** hyperdense clots and **(B)** non-hyperdense clots, and by size category: **(C)** small (0–<50 mm³), **(D)** medium (50–<150 mm³), and **(E)** large (≥150 mm³) clots. Each panel shows the fitted linear regression line (red) and reports the corresponding Pearson or Spearman correlation coefficient (chosen based on normality) and p value, illustrating the inverse association between higher uncertainty and lower DSC across clot phenotypes. *Abbreviations: U_clot = clot-level uncertainty score; mm³ = cubic millimeters; DSC = dice similarity coefficient.*

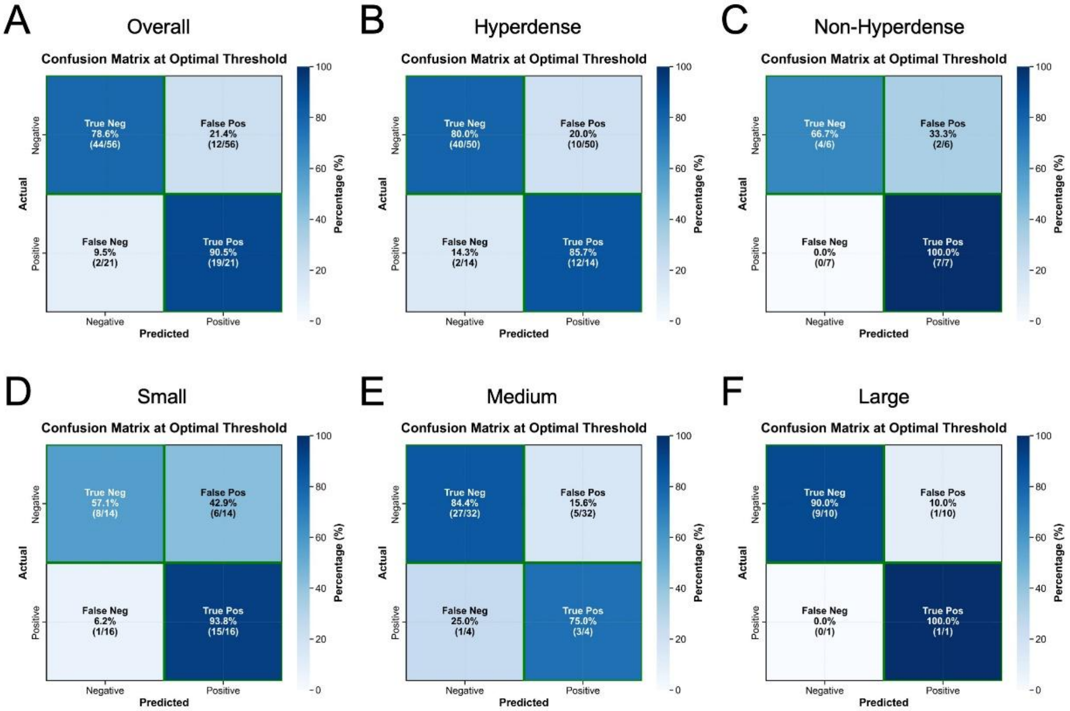


**Supplemental Figure 3. Confusion matrices for uncertainty-based classification of segmentation quality across clot subgroups.** Confusion matrices showing classification of cases as “poorly segmented” (Dice < 0.60, positive) versus “well segmented” (Dice ≥ 0.60, negative) using the clot-level uncertainty score U_clot with the optimal threshold U_clot = 0.323. Panels show results for **(A)** the overall cohort, **(B)** hyperdense clots, **(C)** non-hyperdense clots, and by size category: **(D)** small (0–<50 $mm^3$), **(E)** medium (50–<150 $mm^3$), and **(F)** large (≥150 $mm^3$). Each matrix summarizes the number of true negatives, false positives, false negatives, and true positives for U_clot-based triage within the corresponding subgroup. *Abbreviations: Neg = negative; Pos = positive;* $mm^3$ *= cubic millimeters.*

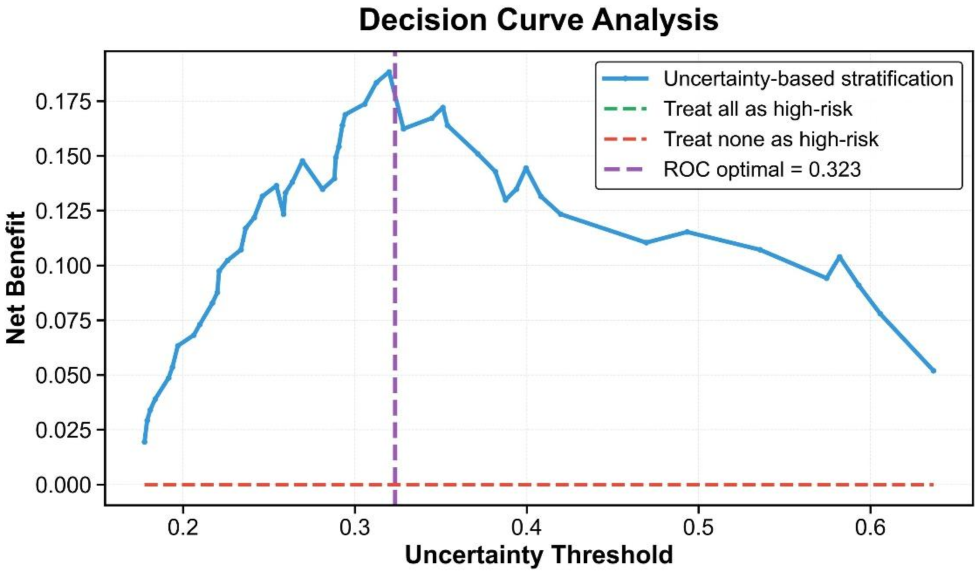


**Supplemental Figure 4. Decision-curve analysis for U_clot-based triage of clot segmentations.** Decision-curve analysis evaluating the clinical utility of using the clot-level uncertainty score U_clot to flag poorly segmented cases (Dice < 0.60) for manual review. Net benefit is plotted as a function of the decision threshold, comparing three strategies: reviewing no segmentations ("treat none"), reviewing all segmentations ("treat all"), and U_clot-guided triage. The U_clot-based strategy yields positive net benefit across a broad range of thresholds and exceeds both "treat all" and "treat none," with a maximal net benefit of 0.188 at the threshold corresponding to the ROC-derived optimal cut-off U_clot = 0.323, equivalent to correctly flagging ~19 additional poorly segmented cases per 100 patients after accounting for unnecessary reviews. *Abbreviations: U_clot = clot-level uncertainty score; ROC = receiver operating characteristic.*

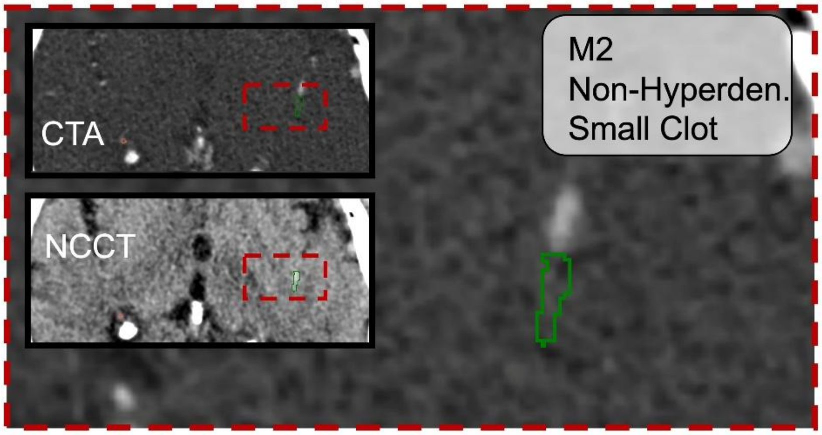


**Supplemental Figure 5. Example of a missed thrombus segmentation.** Representative case illustrating a failure mode of the model. The main panel shows a zoomed-in CTA slice with the manually segmented thrombus (ground truth, GT; green) in the M2 segment of the middle cerebral artery (distal to the MCA bifurcation). Picture-in-picture views display the corresponding zoomed-out CTA and NCCT for anatomical context. This thrombus was non-hyperdense (median NCCT HU < 51) and small (0–50 $mm^3$). No predicted clot mask is shown because the model failed to generate any overlapping segmentation for this case. *Abbreviations: CTA = CT angiography; NCCT = non-contrast CT; GT = ground truth; MCA = middle cerebral artery; M2 = second segment of MCA.*
